# Assessment of Acute Stroke Care, Metrics and Outcomes: Analysis from the Pre-implementation Phase of the IMPETUS Study

**DOI:** 10.1101/2025.06.19.25329605

**Authors:** Shweta Gupta, Rohit Bhatia, Padma Srivastava, Partha Haldar, Mamta Bhushan Singh, Manish Salunkhe, Imnameren Longkumer, Deepshikha Prasad, Risha Sarkar, Vijay Sardana, Dilip Maheshwari, Bharat Bhushan, Alok Verma, Nikhil Dongre, Nikhil Sahu, Samhita Panda, Sucharita Anand, Biman Kanti Ray, Inder Puri, Paresh Zanzmera, Amit Gamit, Sanjeev Kumar Bhoi, Menka Jha, Priyanka Samal, Seepana Gopi, Garuda Butchi Raju, Amit Bhardwaj, Raminder Singh Sibia, Rupinderjeet Kaur, Ashutosh Tiwari, Niraj Kumar, Mritunjai Singh, Kiran Bala, Surekha Dabla, M.P.S Chawla, Jyoti Garg, Shishir Chandan, Rupali Malik, Thomas Iype, P Chithra, Ashok Kumar, Abhay Ranjan, Ravinder Garg, Sulena Sulena, Pramod Darole, Gurpreet Chhina, Shalin Shah, Sudhir Shah, Gajendra Ranga, Smita Nath, Alvee Saluja, L H Ghotekar, Venugopalan Vishnu, Roopa Rajan, Anu Gupta, Deepti Vibha, Rajesh Singh, Awadh Kishor Pandit, Ayush Agarwal, Amit Rohila, Pushpinder Khera, Sarbesh Tiwari, S Bhaskar, Mayank Garg, Anupam Dey, Satyabarta Guru, Suprava Naik, T. Sateesh Kumar, Minakshi Dhar, Naman Agarwal, Mayank Patel, Pranav Joshi

## Abstract

**Background:** Stroke is a major cause of death and disability in India. Despite national programs and guidelines, stroke care continues to face challenges. Many stroke patients seek care at government medical colleges. To date, there are no studies from India that comprehensively assessed the quality of acute stroke care. This study aims to evaluate acute stroke care in its pre-implementation phase across 22 medical colleges using key indicators for optimal care.

**Methods:** IMPETUS stroke is a multicentric, prospective, multiphase, mixed-methods, quasi-experimental implementation study, comprising three phases, initiated in October 2021. During its pre-implementation phase, a baseline assessment of the existing components of stroke care was performed using pre-structured case report form, among prospectively enrolled acute stroke patients at 22 medical colleges.

**Results:** A total of 2,018 patients were enrolled during the pre-implementation phase. Mean (SD) age was 59 (14) years with male preponderance (64%); 69% had an onset <24 hours, majority had ischemic stroke (60%), followed by ICH (38%). Key risk factors were hypertension (80%), diabetes (30%), smoking (22%), alcohol abuse (24%) and previous stroke (21%). Imaging performed: CT (69%), CTA (18%) and MRA (14%). Intravenous thrombolysis was administered in 39% eligible, predominantly with TPA (72%). In-hospital delay was the most common reason for not receiving thrombolysis (44%). The status of stroke time metrics (in minutes) was: onset-to-door 660 (IQR 285–1682), door-to-CT 95 (IQR 46–274), onset-to-needle 201 (165-250), CT-to-needle 36 (23-50), and door-to-needle time 67 (48-90). Other important stroke care indices were also evaluated. In-hospital mortality was 19% and 33% of patients achieved modified Rankin scale score 0–2 at 90-days.

**Conclusion:** These comprehensive data provide a representative baseline status of acute stroke care in India, which will be useful in comparing advancements of stroke care during the implementation phase of the study and improve policy making.

## Introduction

Worldwide, stroke has emerged as the second leading cause of death and the third leading cause of combined death and disability (as expressed by disability-adjusted life-years lost-DALYs) according to the most recent Global Burden of Disease (GBD) 2021 estimates.^1^ There was a substantial increase in the incidence of stroke with low-income and low-middle-income countries (LMICs) carrying the largest share of the global stroke burden. India accounts for 13.3% of the global disability-adjusted life years (DALYs) lost due to stroke with a relatively younger age of onset compared to the Western population.^2^

Stroke care is not uniform across both public and private sector and could be related to several factors including felt need, infrastructural deficiency, limited trained or experienced manpower and administrative support.^3^ While a national program for stroke^4^ and established stroke management guidelines exists^5^, the delivery of organized stroke care continues to face significant challenges in India. Medical colleges, as integral components of the public health system, functions as essential links between rural, district, and other tertiary levels of care centres.^6^ However, in the absence of a systematic performance evaluations, the quality and outcomes of stroke care provided are often presumed to be optimal, rather than being substantiated by empirical evidence. To date, there are no studies from India that comprehensively assessed the quality of acute stroke care.

The implementation of an evaluation and treatment package for uniform stroke care across medical colleges of India (IMPETUS stroke) study^3^ was an implementation research study conducted between October 2021 to December 2024 with the aim to improve stroke care across 22 medical colleges from the time of stroke recognition in the emergency, in-patient management (admitted patients), secondary stroke prevention, and appropriate discharge planning. This study is one of the first prospective, multicenter evaluations focused specifically on public-sector tertiary medical colleges. It provides a detailed, real-world snapshot of stroke care across a wide range of settings before the implementation of any standardized intervention. The present study outlines the status of stroke care observed during the pre-implementation phase of the IMPETUS stroke study across all the collaborating centres.

## Methods

### Aim and objective

The objective of the present study was to observe the status of stroke care across the 22 collaborating centres during the pre-implementation phase of the IMPETUS stroke study, as assessed through various indicators that are essential for optimum stroke care.

### Study Design

IMPETUS stroke was a multicentric, prospective, multiphase, mixed-methods, quasi-experimental implementation study intended to examine the changes in a select set of acute stroke care-related indicators over time within the sites exposed to the same implementation strategy. It comprised three phases: Phase I (pre-implementation), Phase II (implementation) and Phase III (post implementation). The study was initiated in October 2021. Phase 1 is the pre-implementation phase which was expanded from a period of 3 to 5 five months duration, wherein a baseline survey assessment of the existing components of stroke care was performed. Baseline quantitative data was collected on patients using a predefined structured case record form to assess existing status of functioning for the following components: acute stroke evaluation and treatment, risk factor assessment, in-hospital care, discharge planning and secondary prevention, rehabilitation, and caregiver education. The respondent was the key informant at the site level out of the personnel engaged in stroke care (Staff, doctors, and nurses). For each of the patients recruited during this phase, an outcome at the end of 3 months was abstracted from medical records & telecommunication or in person follow up. The data collected has been analyzed on varied parameters using time-series analysis During Phase 1, focused group discussions were held to understand barriers and facilitators of stroke care. This is followed by the implementation phase where main intervention is periodic education and training of the staff, meetings with the site administrators, and education and training of caregivers. Following this implementation phase, data collection is continued to observe the level of implementation and sustainability. The outcomes of implementation shall be assessed on defined indicators of stroke care, mortality, and disability.

### Study Settings

The study was conducted in the collaborating 22 medical colleges and hospitals stretched across 14 different cities from 12 different states of India. All medical colleges are tertiary medical centers situated either in the district headquarters or the state capitals. The Institutional Ethics Committee of each collaborating centers approved the study prior to commencement.

### Study Participants

All consecutive patients with acute stroke (ischemic stroke, intracerebral hemorrhage and cerebral venous thrombosis) admitted in the emergency and inpatients units with suspected acute stroke up to 72 h of onset presenting to the emergency of the collaborating institutions of the study period were recruited after an informed consent was obtained. Caregivers of patients with stroke that were defined as any family member/relative who identifies themselves as a caregiver and spends at least 6 hours per day with the patient were also recruited. Paid professional caregivers were not eligible for the study participation.

### Study Tool

The authors designed the patient information sheet and patient informed consent form which outlines the purpose of the project and maintenance of confidentiality of the patient information. A predefined checklist was used in which nine correlated forms attempted to understand the treatment of patients at the 22 selected medical colleges. The following information was collected: 1) Baseline admission details 2) Thrombolysis & thrombectomy data 3) Laboratory details 4) In hospital details upto 72 hours 4) Discharge details 5) Caregiver Knowledge assessment 6) Follow up information. This extensive case record forms quantitatively assess different parameters of patient care, emergency, in hospital management, laboratory details and radio-diagnostic facility. It also captures the availability of different resources for in-hospital management, discharge planning and rehabilitation. The Redcap database was used to enter the details by each collaborating site.

### Study Outcomes

The present study assessed the current status of stroke care on various parameters observed during a specific period of the IMPETUS stroke study. Three months patient disability outcome was assessed on modified Rankin score dichotomized at 2 or below as a good outcome.

### Statistical Analysis

Descriptive statistics were employed to summarize the patient’s demographic and clinical characteristics, with data presented as mean (standard deviation), median (interquartile range) or frequency (percentage), as appropriate. Patient’s demographic and admission characteristics were stratified according to the type of stroke (ischemic stroke, intracerebral hemorrhage and cerebral venous thrombosis). Differences between these stratified groups were assessed using chi square test for categorical variables and student’s t test for continuous variables. A p-value <0.05 was considered statistically significant. STATA version 14.1 was used to perform all data analyses.

## Results

### Demographic Characteristics

Overall, a total of 2018 patients were recruited during the pre-implementation phase of the study across all the participating medical centres with a mean (SD) age of 59 (14) years. Among these, 64% were males and 36% were females. Patients presenting with an onset of symptoms within 24 hours comprised 69% whereas 18% and 13% of patients were reported with a symptom onset within 24-48 and 48-72 hours, respectively. Majority of the patients were presented with Ischemic stroke (60%), followed by Hemorrhagic stroke (38%) and cerebral sinovenous thrombosis (2%). The National Institute of Health Stroke Scale (NIHSS) at admission was only recorded in 20% of patients. Vital monitoring in emergencies such as blood pressure and blood sugar were measured in 97% and 63% respectively (Table 1). Hypertension was the most common and major risk factor observed in 80% of the patients. Other risk factor includes diabetes (30%), smoking (22%), alcohol consumption (24%), coronary artery heart disease (9%), rheumatic heart disease (3%), atrial fibrillation (5%) and dyslipidemia (4%), previous stroke (21%) and family history of stroke in 13% of the cases (Table 1).

**Table 1.**
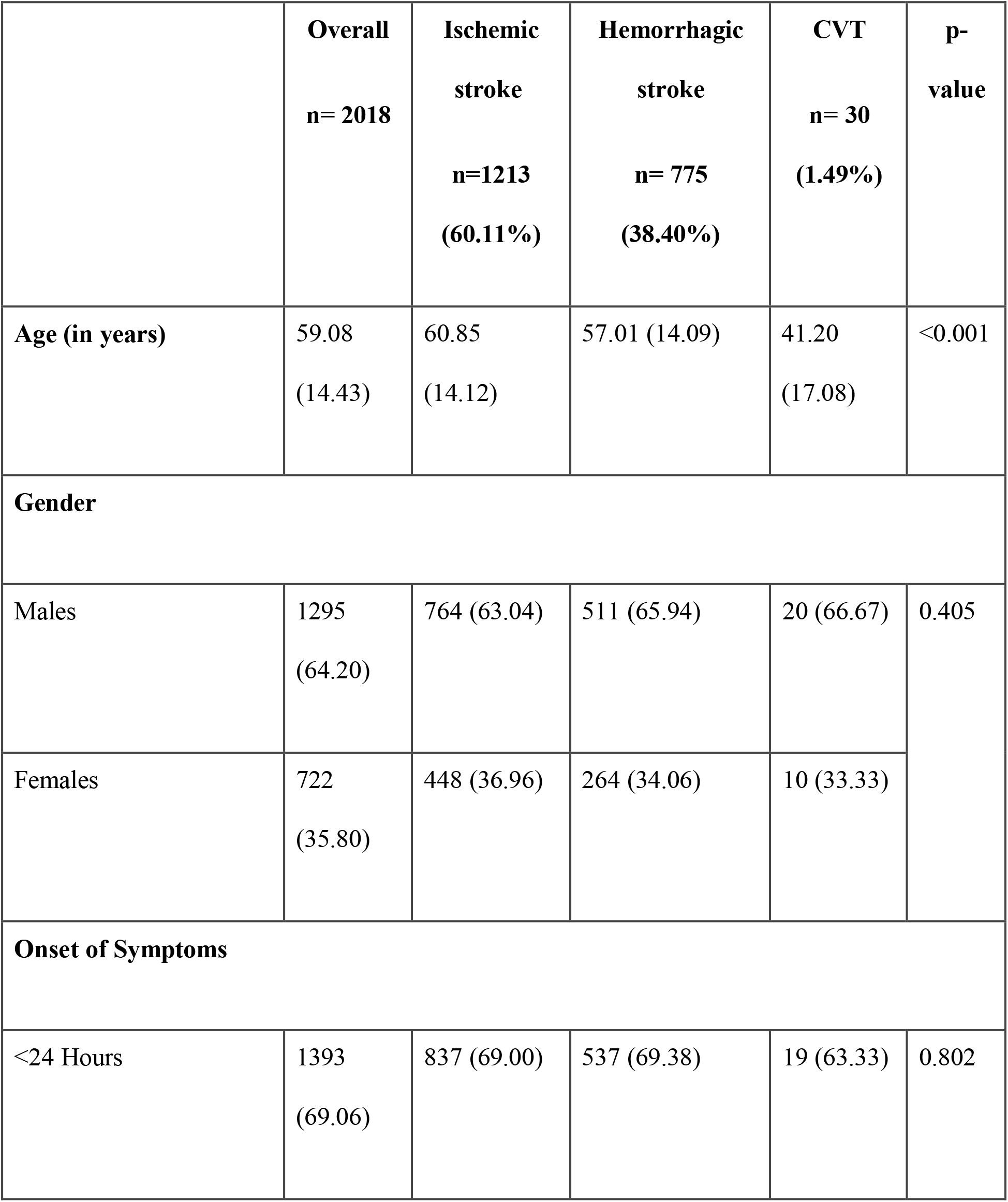
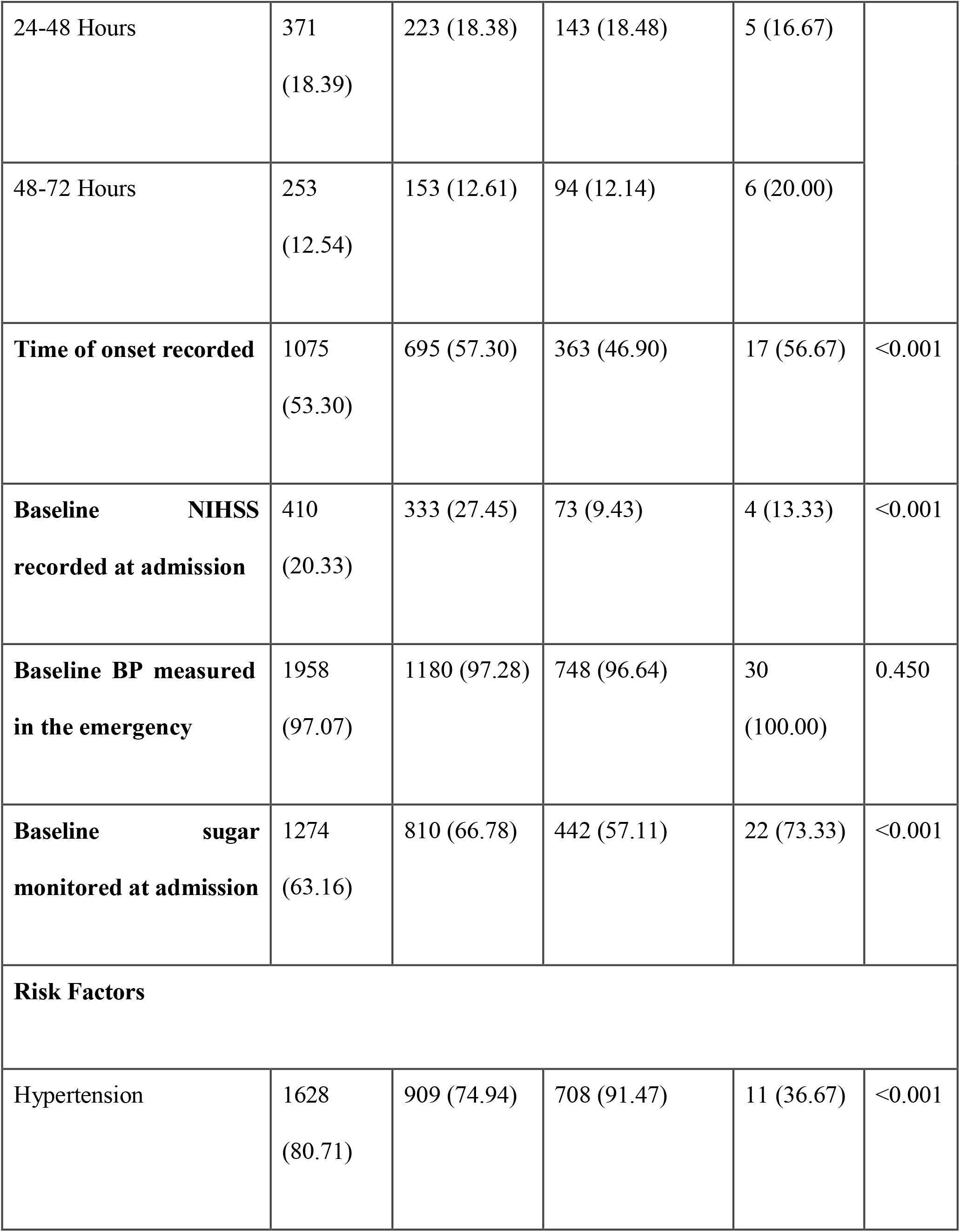
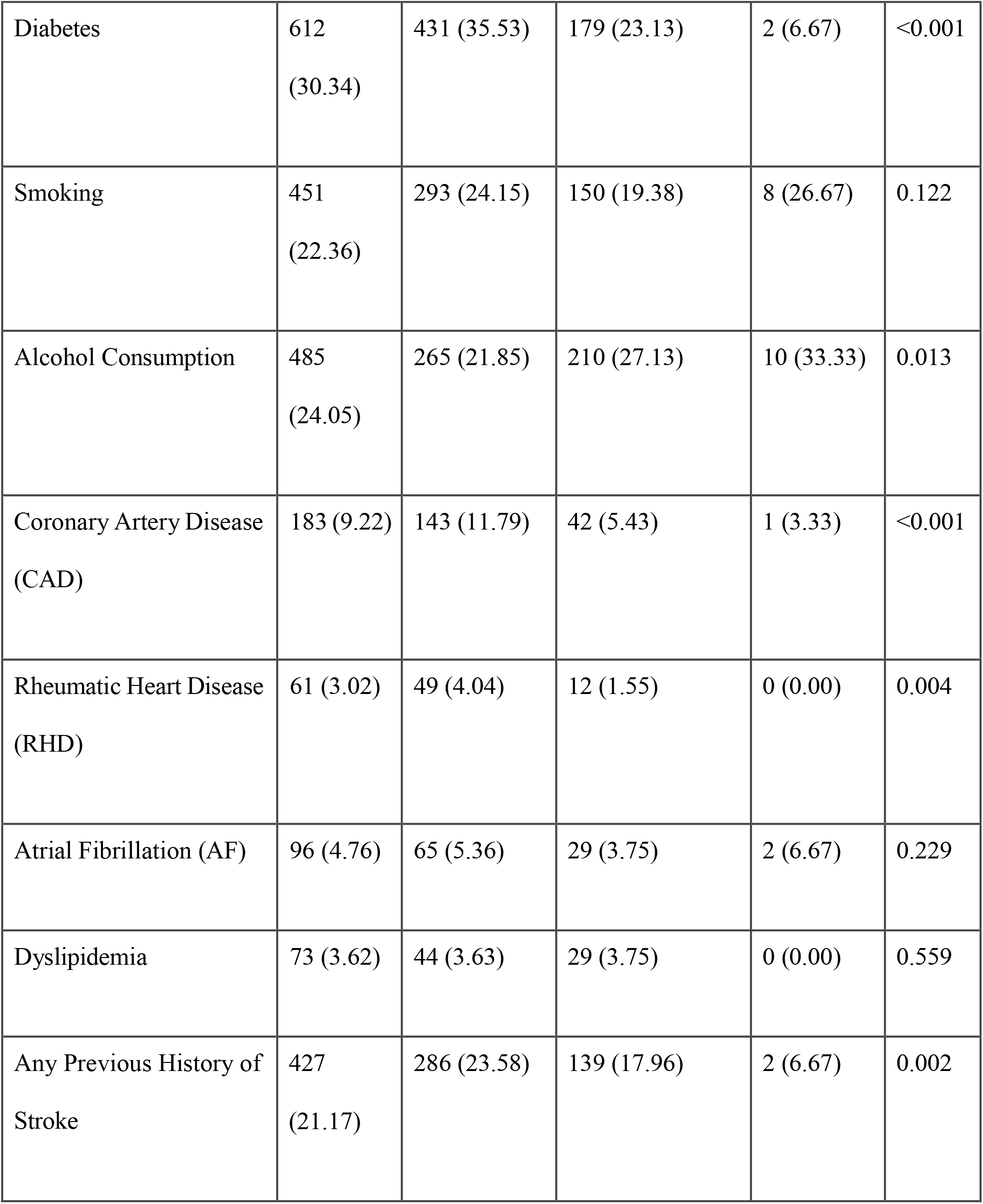
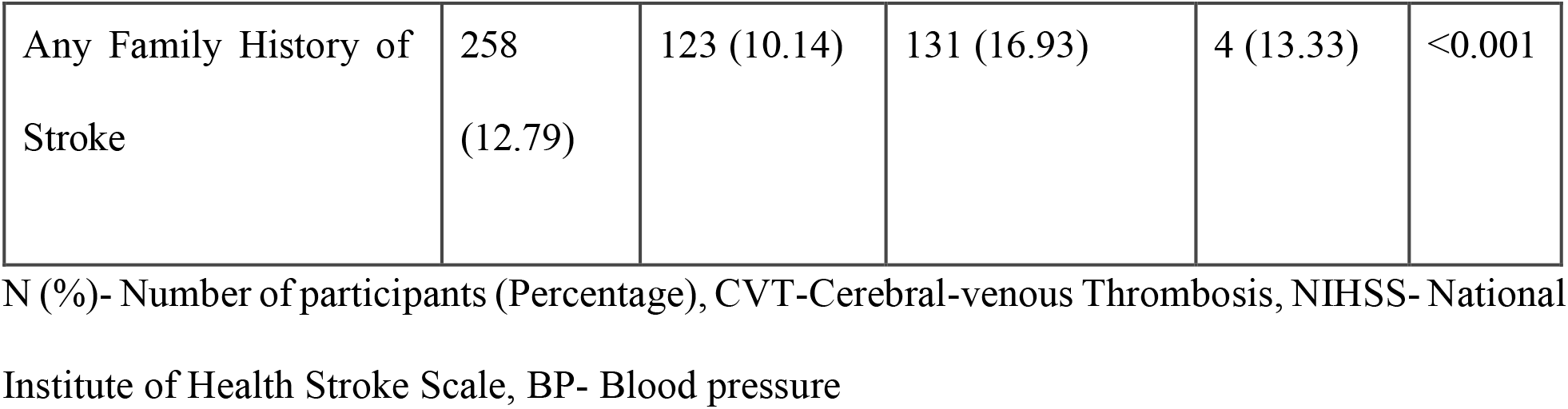
Admission characteristics of admitted patients.

|  | Overall<br>n= 2018 | Ischemic<br>stroke<br>n=1213<br>(60.11%) | Hemorrhagic<br>stroke<br>n= 775<br>(38.40%) | CVT<br>n= 30<br>(1.49%) | p-value |
| --- | --- | --- | --- | --- | --- |
| Age (in years) | 59.08<br>(14.43) | 60.85<br>(14.12) | 57.01 (14.09) | 41.20<br>(17.08) | <0.001 |
| Gender |  |  |  |  |  |
| Males | 1295<br>(64.20) | 764 (63.04) | 511 (65.94) | 20 (66.67) | 0.405 |
| Females | 722<br>(35.80) | 448 (36.96) | 264 (34.06) | 10 (33.33) |  |
| Onset of Symptoms |  |  |  |  |  |
| <24 Hours | 1393<br>(69.06) | 837 (69.00) | 537 (69.38) | 19 (63.33) | 0.802 |
| 24-48 Hours | 371<br>(18.39) | 223 (18.38) | 143 (18.48) | 5 (16.67) |  |
| 48-72 Hours | 253<br>(12.54) | 153 (12.61) | 94 (12.14) | 6 (20.00) |  |
| **Time of onset recorded** | 1075<br>(53.30) | 695 (57.30) | 363 (46.90) | 17 (56.67) | <0.001 |
| **Baseline NIHSS recorded at admission** | 410<br>(20.33) | 333 (27.45) | 73 (9.43) | 4 (13.33) | <0.001 |
| **Baseline BP measured in the emergency** | 1958<br>(97.07) | 1180 (97.28) | 748 (96.64) | 30<br>(100.00) | 0.450 |
| **Baseline sugar monitored at admission** | 1274<br>(63.16) | 810 (66.78) | 442 (57.11) | 22 (73.33) | <0.001 |
| **Risk Factors** |  |  |  |  |  |
| Hypertension | 1628<br>(80.71) | 909 (74.94) | 708 (91.47) | 11 (36.67) | <0.001 |
| Diabetes | 612<br>(30.34) | 431 (35.53) | 179 (23.13) | 2 (6.67) | <0.001 |
| Smoking | 451<br>(22.36) | 293 (24.15) | 150 (19.38) | 8 (26.67) | 0.122 |
| Alcohol Consumption | 485<br>(24.05) | 265 (21.85) | 210 (27.13) | 10 (33.33) | 0.013 |
| Coronary Artery Disease<br>(CAD) | 183 (9.22) | 143 (11.79) | 42 (5.43) | 1 (3.33) | <0.001 |
| Rheumatic Heart Disease<br>(RHD) | 61 (3.02) | 49 (4.04) | 12 (1.55) | 0 (0.00) | 0.004 |
| Atrial Fibrillation (AF) | 96 (4.76) | 65 (5.36) | 29 (3.75) | 2 (6.67) | 0.229 |
| Dyslipidemia | 73 (3.62) | 44 (3.63) | 29 (3.75) | 0 (0.00) | 0.559 |
| Any Previous History of<br>Stroke | 427<br>(21.17) | 286 (23.58) | 139 (17.96) | 2 (6.67) | 0.002 |
| Any Family History of Stroke | 258<br>(12.79) | 123 (10.14) | 131 (16.93) | 4 (13.33) | <0.001 |

N (%) - Number of participants (Percentage), CVT-Cerebral-venous Thrombosis, NIHSS- National Institute of Health Stroke Scale, BP- Blood pressure

### Imaging and Acute stroke treatment

Non-contrast computed tomography (NCCT) was performed in the majority of the medical colleges (69%) at admission (Table 2). In many centres, other imaging facilities such as CT Angiography (73%), MR Angiography (73%) and Doppler neck vessels (53%) were not performed when indicated due to a number of varied reasons.

**Table 2:** Imaging characteristics of admitted patients.

|  | **n (%)** |
| --- | --- |
| **NCCT at admission** |  |
| a. Yes | 1406 (69.67) |
| b. No | 154 (7.63) |
| c. Done outside of study centre | 458 (22.70) |
| **CT Angiography** |  |
| a. Indicated | 1431 (70.95) |
| b. Not indicated | 586 (29.05) |
| **CT Angiography done among indicated patients** |  |
| a. Yes | 367 (25.65) |
| b. No | 1052 (73.51) |
| c. Not available | 12 (0.84) |
| **MR Angiography** |  |
| a. Indicated | 1188 (58.90) |
| b. Not indicated | 829 (41.10) |
| **MR Angiography done among indicated patients** |  |
| a. Yes | 293 (24.66) |
| b. No | 874 (73.57) |
| c. Not available | 21 (1.78) |
| **Doppler Neck vessels** |  |
| a. Indicated | 978 (48.51) |
| b. Not indicated | 1038 (51.49) |
| **Doppler Neck vessels done among indicated patients** |  |
| a. Yes | 420 (42.94) |
| b. No | 518 (52.97) |
| c. Not available | 40 (4.09) |

N (%) - Number of participants (Percentage), NCCT - non-contrast computed tomography, CT - computed tomography, MR - magnetic resonance

Among ischemic stroke patients eligible for thrombolysis (16%), intravenous thrombolysis (IVT) was administered in 39% of patients. Tissue plasminogen activator (TPA) was the most common agent used for IVT (Table 3). In-hospital delay after admission (45%) was one of the major reasons for patients not being thrombolysed (Table 3) followed by IVT non availability (28%), minor stroke (12%), negative consent (9%) and non-affordability (6%). Endovascular therapy (EVT) was provided to only 1% of patients because in most of the centres EVT services were not available (33%). Several other reasons for exclusion are mentioned in Table 3. Time-sensitive quality measures showed that the median duration from symptom onset to hospital admission was 660 minutes (IQR 285–1682). The overall median door-to-CT scan time was 95 minutes (IQR 46– 274), with ischemic stroke patients having a shorter imaging time of 87 minutes (IQR not reported or unclear) compared to 120 minutes (IQR 53–369) for patients with intracerebral hemorrhage (ICH).

**Table 3:**
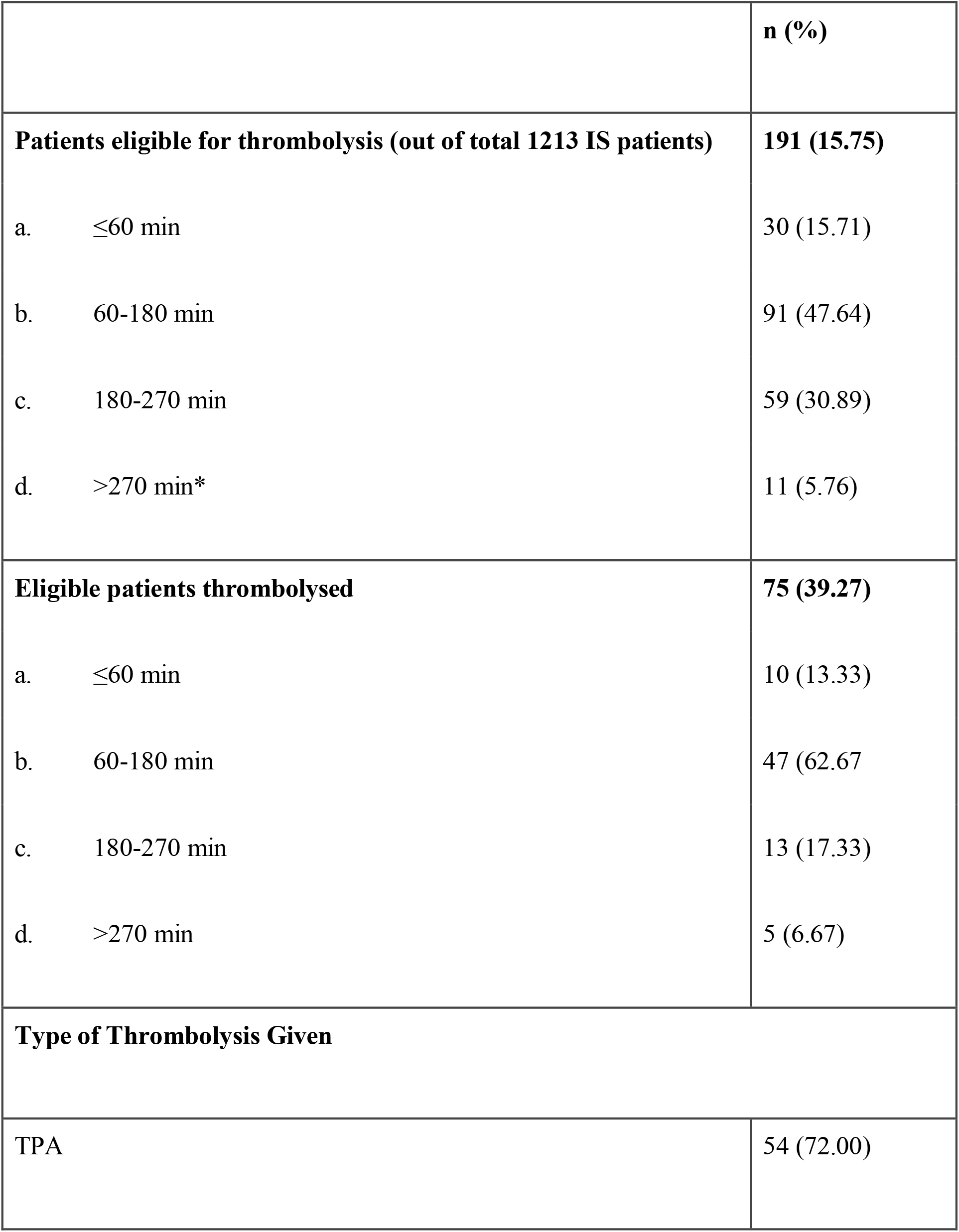
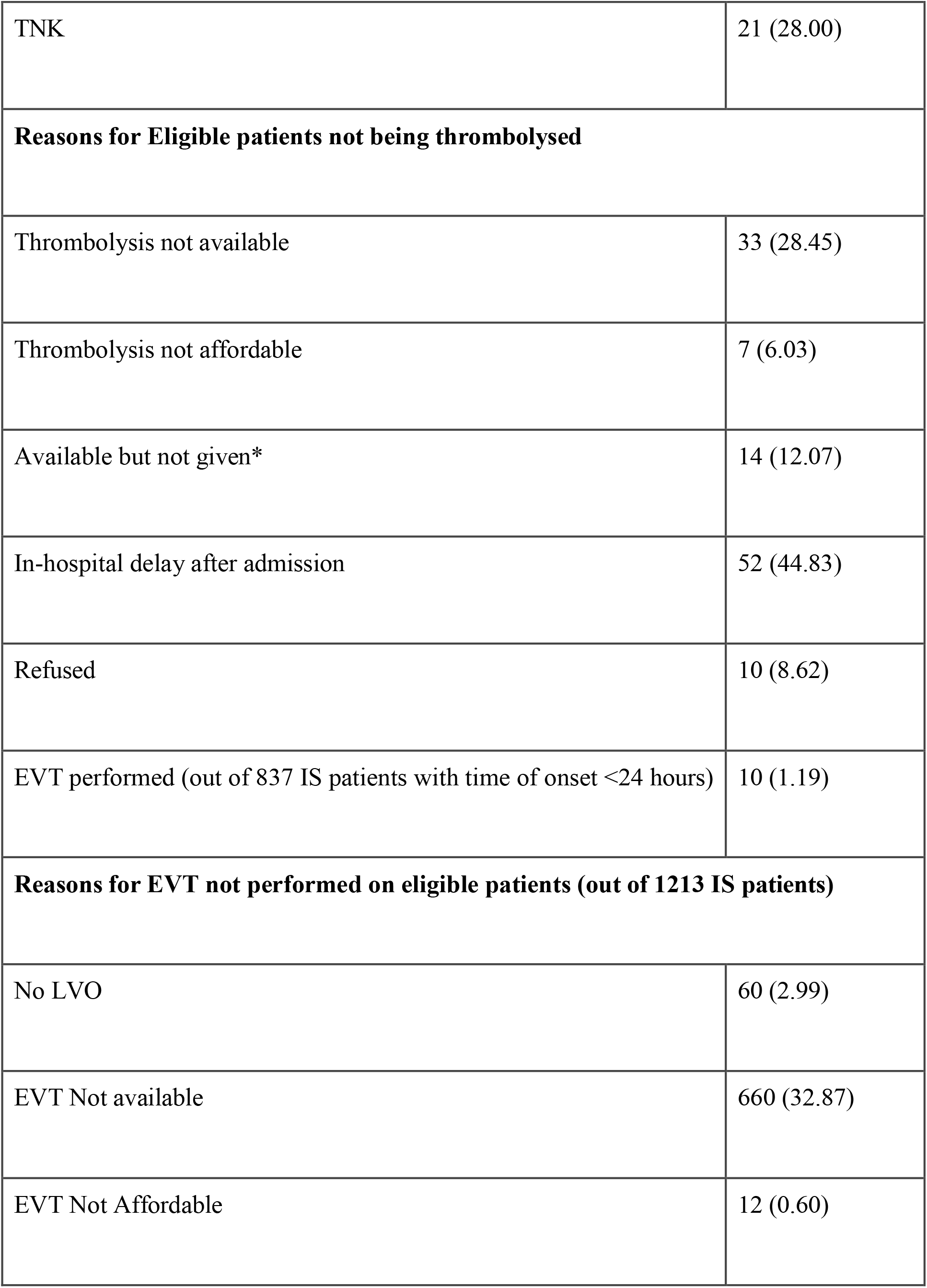
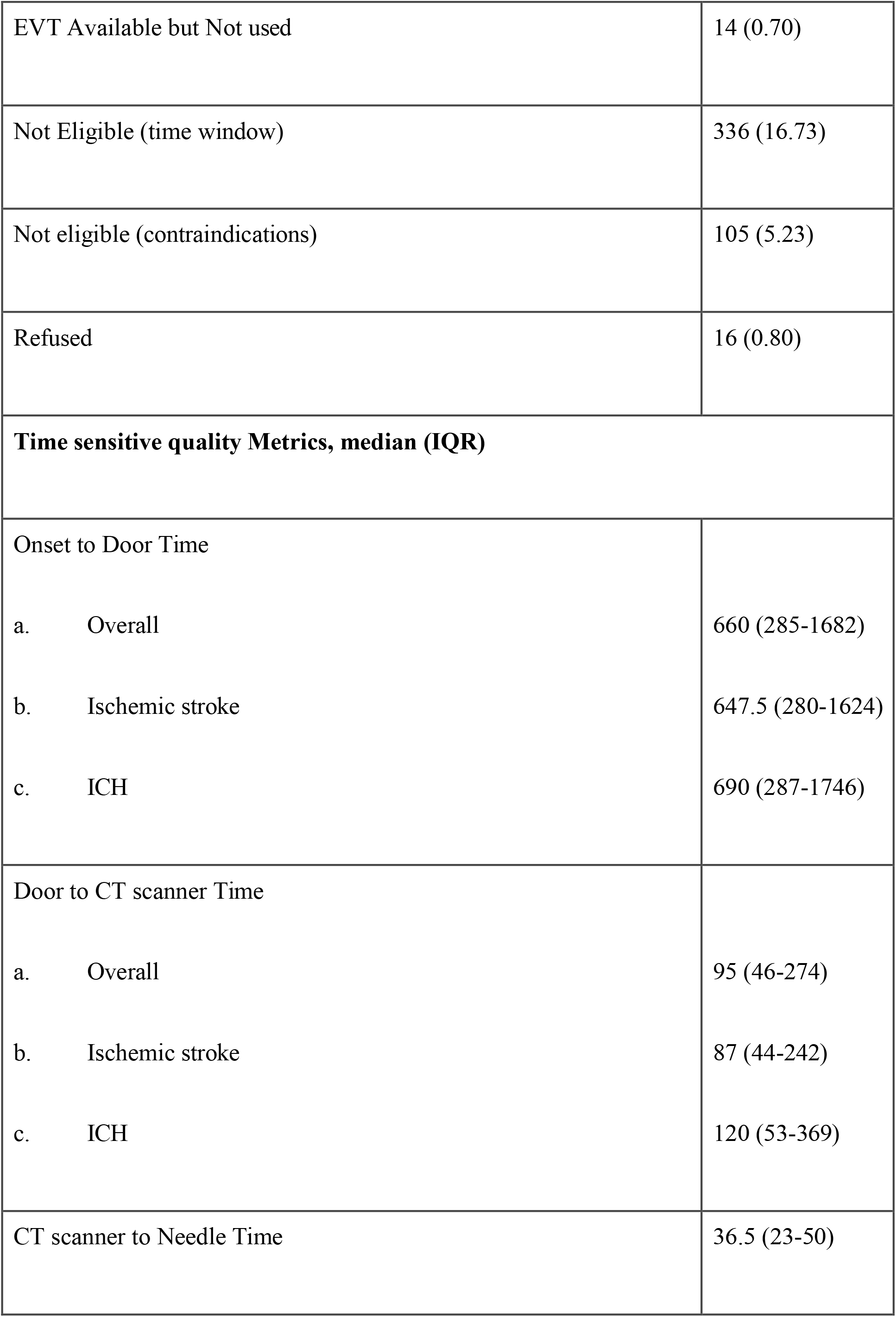
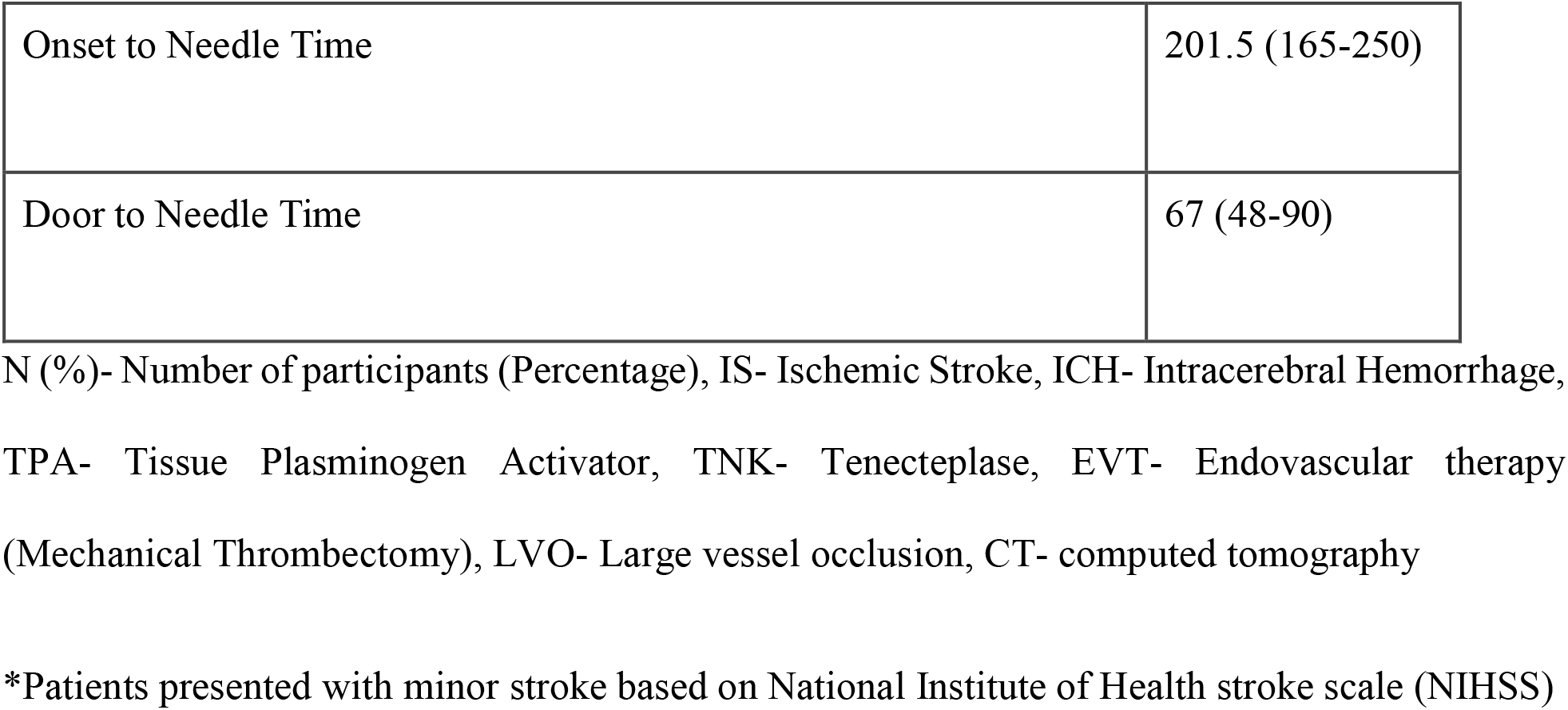
Acute management and stroke metrics among Ischemic Stroke patients.

Among the thrombolysis eligible stroke patients, the imaging time to thrombolysis time was 36.5 (IQR 23-50) minutes, onset of symptoms to thrombolysis time was 201.5 (IQR 165-250) minutes and admission time to thrombolysis time was 67 (IQR 48-90) minutes (Table 3).

### In Hospital Stroke Care Management

During hospital admission, patient level information for three consecutive days (upto 72 hours) was collected to correlate stroke care practices with the patient outcomes. Body temperature and blood pressure were routinely assessed in all centres as compared to level of consciousness (GCS) and sugar monitoring (Table 4). Routine swallow assessments were performed in 55%, 39% and 41% at day 1, 2 and day 3 respectively. It was observed that in 44% of patients, routine assessments were not done at day 1 and around 40% were marked as non-eligible, based upon their level of consciousness. As per the patient’s mobility status, deep vein thrombosis (DVT) prophylaxis was provided to 20% at day 1, not indicated in majority (54%) and 79% did not receive it among those eligible patients. DVT pump (50%) was most commonly used, followed by heparin (41%) and a combination of both (9%). Further details of day 2 and day 3 are mentioned in Table 4. Physiotherapy consultation was done in 56%, 69% and 74% on day 1, 2, and 3, respectively. At centres where rehabilitation personnels were not available or inadequate, physiotherapy consultation was not provided to eligible patients (43%, 30% and 25%, respectively). Similarly, air mattress was not provided to 39% at day 1 due to its unavailability and was only given to 37%, 40% and 42% on consecutive days. However, most of the patients received appropriate positioning (70%, 75% and 78% at day 1, 2 and 3, respectively) during the hospital stay. Stroke care advice was provided to caregivers of the patients (70% at day 3). Further, complications were observed to occur in less proportions (4%, 6% and 7% each day).

**Table 4.** In-Hospital Stroke Care Components.

|  | Day-1* | Day-2* | Day-3* |
| --- | --- | --- | --- |
|  | n = 2006 | n= 1822 | n = 1659 |
|  | n (%) |  |  |
| Vital monitoring |  |  |  |
| GCS Recording | 1235 (61.57) | 851 (46.71) | 733 (44.18) |
| Temperature monitoring | 1690 (84.25) | 1516 (83.30) | 1379 (83.12) |
| BP monitoring | 1528 (76.17) | 1462 (80.24) | 1307 (78.78) |
| Sugar monitoring | 1053 (52.49) | 805 (44.21) | 693 (41.87) |
| Secondary Prevention |  |  |  |
| Swallow assessment |  |  |  |
| a) Indicated | 1214 (60.61) | 891 (60.61) | 752 (45.33) |
| b) Not indicated | 789 (39.39) | 789 (39.39) | 907 (54.67) |
| Swallow assessment among indicated |  |  |  |
| a. Yes | 675 (55.60)) | 352 (39.51) | 314 (41.75) |
| b. No | 539 (44.40)) | 539 (60.49) | 438 (58.25) |
| Deep Vein Thrombosis (DVT) Prophylaxis |  |  |  |
| a. Indicated | 928 (46.26) | 819 (44.95) | 713 (42.98) |
| b. Not indicated | 1078 (53.74) | 1003 (55.05) | 946 (57.02) |
| DVT Prophylaxis among indicated |  |  |  |
| a. Yes | 194 (20.90) | 207 (11.36) | 204 (28.61) |
| b. No | 734 (79.10) | 612 (33.59) | 509 (71.39) |
| Type of DVT Prophylaxis |  |  |  |
| a. Heparin/LMWH | 80 (41.24) | 79 (38.16) | 76 (37.25) |
| b. DVT Pump | 96 (49.48) | 100 (48.31) | 99 (48.53) |
| c. Both | 18 (9.28) | 28 (13.53) | 29 (14.22) |
| Physiotherapy Consultation |  |  |  |
| a. Indicated | 1640 (81.75) | 1565 (85.89) | 1433 (86.38) |
| b. Not indicated | 366 (18.25) | 257 (14.11) | 226 (13.62) |
| Physiotherapy Consultation among indicated |  |  |  |
| a. Yes | 925 (56.40)) | 1089 (69.59) | 1064 (74.25) |
| b. No | 715 (43.60)) | 476 (30.41) | 369 (25.75) |
| Air mattress |  |  |  |
| a. Indicated | 1504 (74.98) | 1375 (75.47) | 1240 (74.74) |
| b. Not indicated | 502 (25.02) | 447 (24.53) | 419 (25.26) |
| Air Mattress among indicated |  |  |  |
| a. Yes | 562 (37.37) | 559 (40.65) | 530 (42.74) |
| b. No | 344 (22.87) | 307 (22.33) | 269 (21.69) |
| c. Not available | 598 (39.76) | 509 (37.02) | 441 (35.57) |
| Appropriate Positioning |  |  |  |
| a. Indicated | 1687 (84.10) | 1531 (84.07) | 1392 (83.91) |
| b. Not indicated | 319 (15.90) | 290 (15.93) | 267 (16.09) |
| Appropriate Positioning among indicated |  |  |  |
| a. Yes | 1189 (70.48) | 1159 (75.70) | 1099 (78.95) |
| b. No | 498 (29.52) | 372 (24.30) | 293 (21.05) |
| **Caregiver Advice** | 1353 (67.45) | 1266 (69.48) | 1161 (69.98) |
| **In hospital Complications Recorded** | 90 (4.49) | 109 (5.99) | 126 (7.60) |

N (%) - Number of participants (Percentage), GCS-Glasgow coma scale, BP-Blood pressure, LMWH-Low molecular weight heparin
\*As per the duration of the stay in the hospital

### Discharge Status and Secondary Prevention

Overall, in-hospital mortality was observed in 388 (19%) patients. A total of 304 (19%) patients were left / discharged against medical advice (LAMA/DAMA) of those who survived [n= 1612 (80%)]. At the time of discharge, in-hospital complications were recorded in 11% of patients. TOAST criteria for etiological identification were not mentioned in the majority (82%) of ischemic stroke patients. Large artery atherosclerosis (42%) was found to be the most common etiological factor based upon the evaluation (Supplementary Table 1). 43% hypertensive of patients had received the risk factor advice at discharge. Advice for other risk factors such as diabetes and dyslipidemia management was given in 17% and 8%, respectively. Patient care advice regarding tracheostomy (9%), catheter care (22%), Ryle’s Tube feeding (13%) and positioning advice (36%) were mentioned on discharge summary. It was also observed that care advice was not mentioned in proportions of patients when indicated in the discharge summary. Advice on medication adherence (38%), medication dose (98%), timing (91%) and adverse effects (18%) were provided and documented in the discharge summary. 68% caregivers were counselled and properly advised. Follow up advice regarding when to follow up (87%), whom to follow up (62%) and where to follow up (86%) were mentioned on discharge summary.

### mRS at 3 months

Overall, 33% patients achieved a modified Rankin Score (mRs) 0-2 at 90 days follow up and 36%, 26% and 65% in IS, ICH and CVT, respectively (Figure 1 and 2).

**Figure 1:**
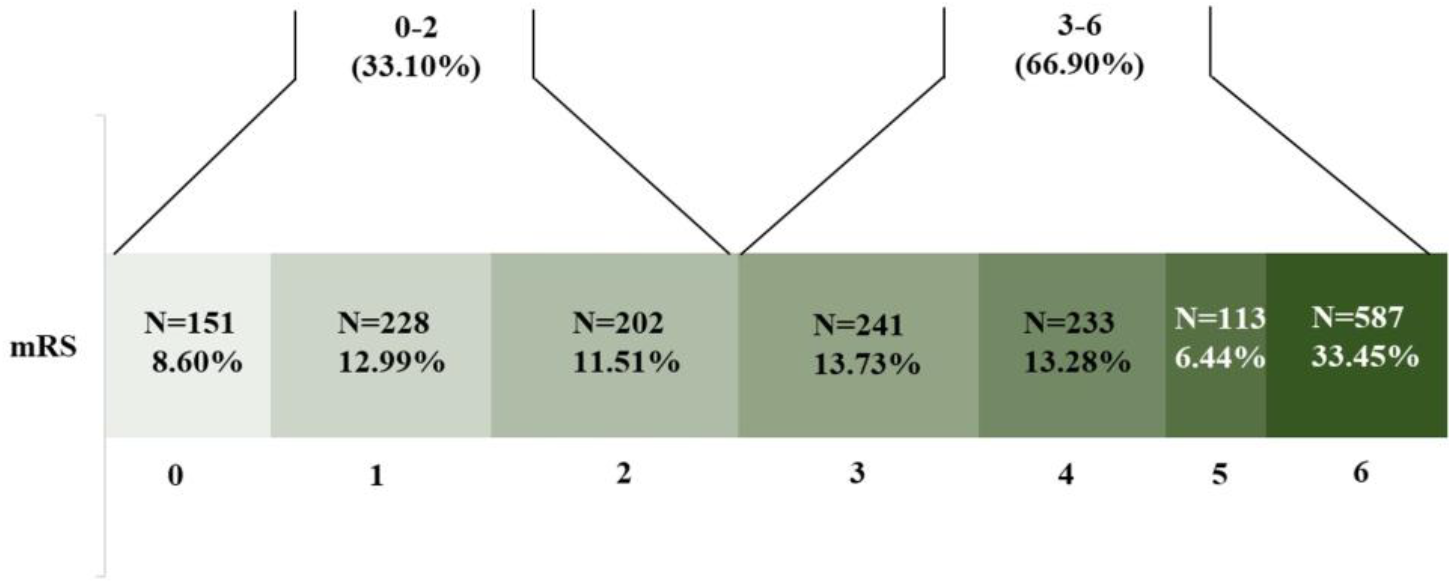
Distribution of modified rankin scale (mRS) scores at 90-days follow-up in the overall cohort, showing the proportion of patients achieving good outcomes (mRS 0-2, 33.10%) versus poor outcomes (mRS 3-6, 66.90%)

**Figure 2:**
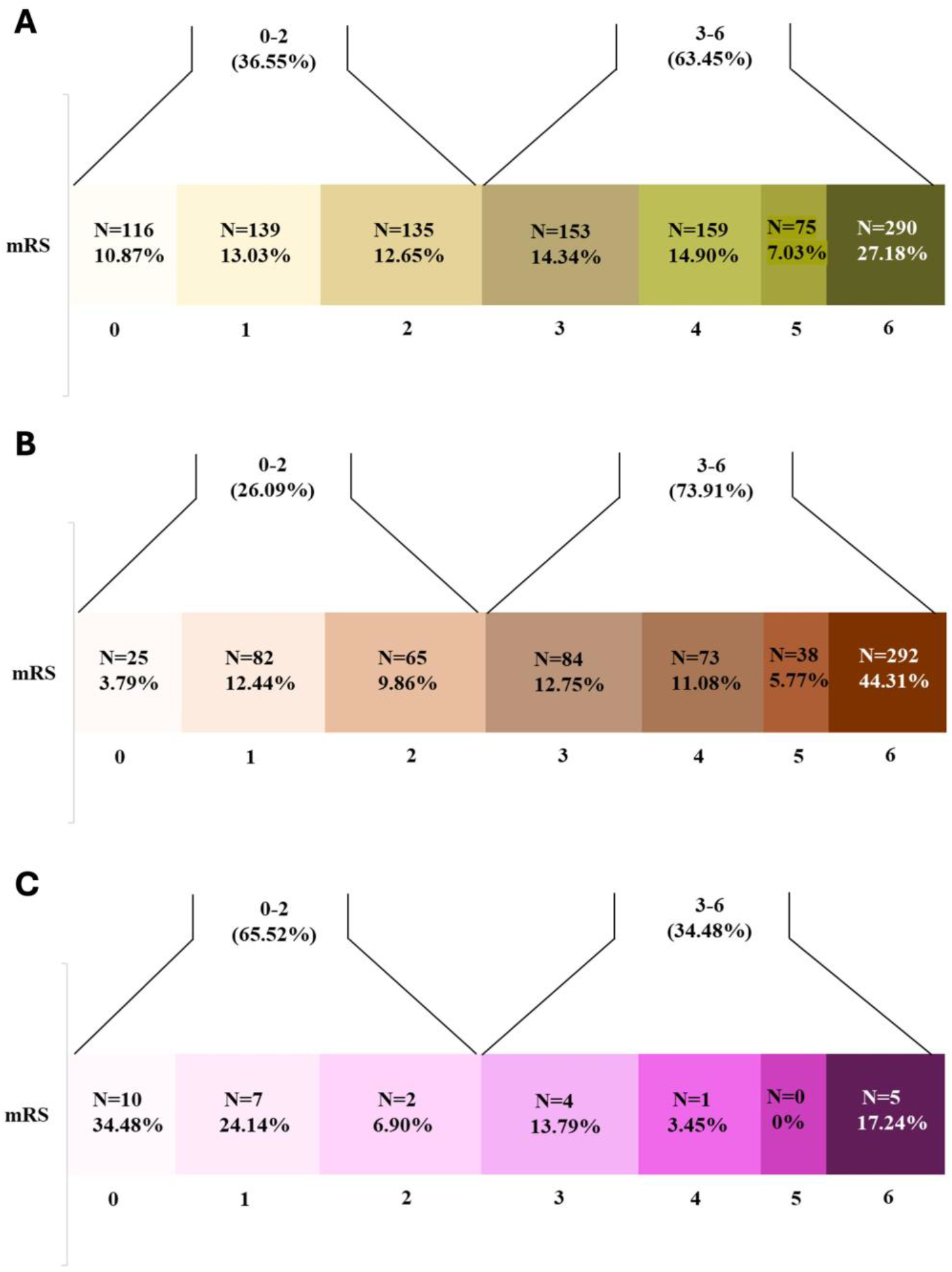
Distribution of modified rankin score (mRS) at 90-days follow up in stroke subtypes. Patients with good (mRS 0-2) versus poor outcomes (mRS 3-6) varied across subtypes: A) Ischemic stroke (36.55% vs 63.45%, respectively); B) Intracranial hemorrhage (26.09% vs 73.91%, respectively); C) Cerebral venous thrombosis (65.52% vs 34.48%, respectively)

## Discussion

The present study highlights the status of stroke care among collaborating centres of the IMPETUS stroke implementation study^3,14^ in its pre implementation phase and the potential gaps in the stroke care pathway. This also helps provide real time information on the various key variables that impact stroke care and thereby its outcome. One of the critical challenges in stroke management is the treatment gap in the public healthcare settings. The available data reflects the status across the stroke care continuum, rapid access, acute stroke treatment and care, discharge and secondary prevention.

Population level studies from LMICs reported higher incidence of stroke and probable increase in the future stroke rates.^7,8^ In the INTERSTROKE study of 12,342 patients with stroke from 108 hospitals in 28 countries, individuals in LMICs more often had severe strokes, intracranial hemorrhages, poorer access to services, and fewer investigations and treatments than those in Higher income countries (HICs).^9^

We observed a large proportion of patients arriving at the hospital after 24 hours of stroke onset. Lack of public awareness about the warning signs of stroke symptoms, sociocultural beliefs and transport accessibility leads to lengthy delays in presenting at the hospital.^10,11^ Timely recognition of stroke onset is crucial for determining a patient’s eligibility for time dependent acute ischemic stroke treatments. Previous study in a tertiary stroke centre from Romania observed that most of the stroke patients arriving after 24 h from onset were living alone and living in rural areas and highlighted the need of stroke awareness program and pre-hospital protocols.^12^

Availability and accessibility of the 24/7 functional CT scan or MRI facility should be mandatory as per the national guidelines for stroke prevention and management.^13^ CT / MR Angiography was performed in a limited proportion of patients. Routine vascular imaging is generally not performed in many centres as surveyed by us previously.^14^ It may be due to issues with availability, manpower, or a general practice pattern. The absence of easy and timely access to brain imaging adds to the difficulties in diagnosing stroke in LMICs. In Ghana, only two-thirds of hospitals have a functional CT scanner available during working hours on weekdays.^15^ A systematic review of stroke services in Africa showed that only 13–36% of patients underwent CT or MRI scans due to its unavailability and financial constraints.^16^ As per guidelines recommended by Healthcare professionals from the Indian Stroke Association, and other International stroke guidelines, stroke centres should be capable of performing imaging within 30 minutes of a patient presenting at the centre.^17^ In the present study, door to CT time exceeded the ideal threshold with an overall median CT time of 97 mins. Single call notification or stroke code can significantly help in rapid evaluation of stroke patients.^18^ The response time towards a patient in the emergency may also vary depending upon the perceived “eligible” patient for thrombolysis and delays could happen due to other sick patients being prioritized in an extremely busy emergency in public hospitals.

NIHSS is the most reliable and commonly used stroke severity tool in clinical trials for the administration of thrombolytic drugs.^19,20^ A small proportion of baseline NIHSS assessment was observed. It is likely due to lack of training, awareness, stroke trained physicians or nurses and overburdened healthcare settings. At most centres, the medicine resident is the first contact, and not a stroke physician. The low rate of NIHSS documentation and delayed imaging likely reflects deeper system-level issues, such as the absence of standardized triage pathways, poor coordination between emergency, radiology, and neurology services, and lack of structured prehospital care. Although regular training and education may reinforce improvement, there are challenges that go beyond individual training and require broader organizational and process-level changes tailored to the Indian public health system.

Intravenous thrombolysis and mechanical thrombectomy are an effective and approved treatment for the acute stroke management, however, their use remains limited in LMICs.^21,22^ The present study observed a small proportion of ischemic stroke patients eligible for thrombolysis and within them, only one third were thrombolysed. In-hospital delay after admission and unavailability of the thrombolytic drug were observed as the most common reasons for eligible patients not being thrombolysed. A prospective study from a tertiary care hospital reported pre-hospital delay (81.5%), crowded emergency (77.7%), financial constraints (76.7%) and delay in CT scan (61.4%) as barriers to thrombolysis.^23^ In Peru, only 2% of patients received thrombolysis due to its unavailability. Similar observations were found for EVT.^24^ In our previous study of infrastructural assessment, EVT services were only available in 27% of the hospitals and only two hospitals were providing it free of cost.^14^ Another survey from Ghana revealed that none of the 11 major hospitals in Ghana were conducting the EVT procedure.^15^ Limited trained professionals, high cost and infrastructural demands limit the use of EVT in LMICs. In a survey conducted by the mission thrombectomy (MT) 2020 plus global network among 75 countries, global mechanical thrombectomy use was poor and LMICs had 88% lower mechanical thrombectomy access when compared to HICs.^25^ Treatments such as thrombolysis and EVT should get support from national and organizational levels to become accessible at low cost or be government funded.

For the implementation of an organized system of stroke care, in-hospital management should have key elements such as stroke units, blood pressure and cardiac monitoring, GCS recording, regular temperature and sugar monitoring, routine swallow assessment and interventions for prevention of secondary complications. Stroke unit admissions is one of the most effective ways to reduce hospital mortality and morbidity but its implementation still remains a challenge in LMICs.^26,27^ In India, there were only 35 stroke units in 2013 with the majority in the private sector.^28^ Infrastructural reorganization to create a designated geographical stroke unit with 4-6 beds and training of healthcare professionals should be emphasized in low resource settings.

The present study found inconsistency in swallow assessment as nearly 27% patients did not undergo swallow evaluation during the first 24 hours despite being eligible. Implementation of standardized protocols for clinical monitoring and management of temperature, sugar and swallowing assessment are highly beneficial.^29^ In a randomized control trial from a tertiary care center from India, reduction in the hospital mortality was reported with a nurses-led fever, sugar and swallowing bundle care.^30^ Data from 64 hospitals from 17 countries across Europe showed improvements in pre-to-post implementation of all three components in both high and low resource settings.^31^ Mandatory implementation of the protocol and frequent training for the nursing officers should be encouraged. Simple interventions such as positioning, provision of air mattress, DVT prophylaxis and mobility assessments can help to reduce hospital complications and improve patient outcomes. Unavailability of the resources and a multidisciplinary team is a major barrier in providing optimum care for stroke patients.

Early mobilization and rehabilitation provide better outcomes for stroke survivors.^17,27^ In this study, physiotherapy consultation was not done for almost one fourth of the patients. Most of the patients received rehabilitation advice as per consultation request. Lack of rehabilitation specialist, policies or guidelines, less prioritization, overburdened wards, lack of dedicated stroke and step-down in-patient rehabilitation units lead to compromised rehabilitation services in hospitals. Among a cohort of 250 stroke patients from Zambia, only 27% patients received physical therapy evaluation during hospitalization. Occupational and Speech therapists were entirely unavailable. Similar findings had been found in mean number of in-hospital physiotherapy session (two sessions per 8-day length of stay in Rwanda, three sessions per 12-day LOS in Tanzania, and two sessions per 7-day LOS in South Africa).^32,33^ During the INTERSTROKE study, it has been found that 77% of patients with stroke in LMICs have moderate to severe functional disability after stroke, compared with 63% in upper-middle-income countries, and 38% in HICs.^9^ In response to the substantial gap between the need for rehabilitation and the capacity of countries to respond to that need, the World Health Organization launched Rehabilitation 2030: A Call to Action.^34^ Family caregivers are an important part of stroke rehabilitation in low-resource settings.^35^ We observed that only a small proportion of caregivers were counselled at the time of discharge for post stroke care. Family led Rehabilitation in India and a nurse-plus-caregiver strategy in Mexico found that task shifting rehabilitation is feasible and does not jeopardize stroke survivors health.^36,37^ Post discharge rehabilitation services in LMICs (31%) are much worse as compared to the HICs (92%).^38^ Incorporating structured educational programs to strengthen caregiver knowledge has proven beneficial in reducing the complications both during hospitalization as well as after discharge at home.^39^ Training sessions using stroke manuals and video modules will be conducted during the implementation phase of this study in order to enhance caregiver knowledge.

Different Stroke models can be adapted to combat these challenges within the healthcare systems of LMICs. Multidisciplinary team care has been identified as a key component in effective stroke care.^40^ In this system, patients with stroke are immediately identified and preferably managed in a separate stroke unit. Training and education sessions for the non-neurologist can be conducted to identify stroke symptoms in the emergency.^41^ Another model that was proven effective is the hub and spoke model for the timely and effective management of the stroke patient.^42^ Studies from India and Brazil have reported on the use of tele-stroke services.^43,,44^ Tele-stroke using smart phone based services can help to address the shortage of neurologists, especially in hard to reach areas and areas in which the population is highly dispersed.^45^

## Strengths and limitations

The strength of study includes its prospective, real world observational data that helps provide insight into the current status of stroke care on various indicators that are essential and needs improvement for optimum stroke care. The current data reflect the existing system of stroke management and can help in the decision making of policies for better stroke care. This can also help to strengthen skilled and trained manpower in medical colleges to impart best available stroke care to patients. Any change or improvement in the stroke care pathway shall be reflected after completion of the implementation and training in the ongoing study. Limitations include data collected from the different medical colleges in urban settings only and economic disparities are likely to exist.

## Conclusion

This study provides the current status of stroke care in different medical colleges in India. The comprehensive data offers a representative baseline status of acute stroke management, which will be useful in comparing implementation assessment and advancements of stroke care. Public awareness programs, implementation of existing guidelines, quality improvement initiatives, increased number of stroke units and stroke ready hospitals, mandatory stroke orientation programs, infrastructural re-organization and capacity building will help in implementation of uniform stroke care pathway and hence improve stroke outcomes.

## Data Availability

Data will be provided on request as per the justification and requirement through email

## Acknowledgments

We acknowledge our funding authority and study participants

